# Best of intent, worst of both worlds: why sequentially combining epidemiological designs does not improve signal detection in vaccine safety surveillance

**DOI:** 10.1101/2022.02.18.22271183

**Authors:** Faaizah Arshad, Martijn J. Schuemie, Evan P. Minty, Thamir M. Alshammari, Lana Y.H. Lai, Talita Duarte-Salles, Stephen Fortin, Fredrik Nyberg, Patrick B. Ryan, George Hripcsak, Daniel Prieto-Alhambra, Marc A. Suchard

## Abstract

**Background:** Vaccine safety surveillance commonly includes a serial testing approach with a sensitive method for “signal generation” and specific method for “signal validation.” Whether serially combining epidemiological designs improves both sensitivity and specificity is unknown.

**Methods:** We assessed the overall performance of serial testing using three administrative claims and one electronic health record database. We compared Type I and II errors before and after empirical calibration for historical comparator, SCCS, and the serial combination of those designs against six vaccine exposure groups with 93 negative control and 279 imputed positive control outcomes.

**Results:** Historical comparator mostly had lower Type II error than SCCS. SCCS had lower Type I error than the historical comparator. Before empirical calibration, serial combination increased specificity and decreased sensitivity. Type II errors mostly exceeded 50%. After empirical calibration, Type I errors returned to nominal; sensitivity was lowest when the methods were combined.

**Conclusion:** We recommend against the serial approach in vaccine safety surveillance. While serial combination produced fewer false positive signals compared to the most specific method, it generated more false negative signals compared to the most sensitive method. Using the noisy historical comparator in front of SCCS deteriorated overall performance in evaluating safety signals.

**Key Messages:**

- Using the serial approach in vaccine safety surveillance did not improve overall performance: specificity increased but sensitivity decreased.
- Without empirical calibration, Type II errors exceeded 50%; after empirical calibration, Type I error rates returned to nominal with negligible change to Type II error rates.
- While prior research has suggested high sensitivity of the historical comparator method in distinguishing true safety signals, there were cases when self-controlled case series was more sensitive.
- Vaccine safety surveillance is becoming increasingly important, so monitoring systems should closely consider the utility and sequence of epidemiological designs.

## 1 Introduction

As of November 2021, over 7 billion doses of coronavirus disease 2019 (COVID-19) vaccines have been administered worldwide [1] [2], with 550 million in the United States (US) [3] [4] and 600 million in Europe [5]. While the rapid, global increase in vaccine uptake brings promise that the pandemic will end soon, undesirable and sometimes life-threatening adverse events can occur after immunization. Public health experts are employing post-vaccine safety surveillance to identify risk factors and complications of vaccine use.

Typically, in vaccine development, companies in the US are only permitted to market their vaccine if approved by the US Food and Drug Administration (FDA). However, due to the COVID-19 emergency, the US FDA granted emergency use authorizations to permit vaccine distribution ahead of the typical approval process. This underscores the need for post-vaccine monitoring to ensure that unapproved vaccines in widespread use are safe.

Additionally, only a limited number of people participate in clinical trials (which are often highly restricted, such as with pregnant women or persons with specific comorbidities like cancer, etc.), so initial data on rare adverse events can be limited and ungeneralizable to real-world populations [6]. After the vaccine is marketed and provided to larger and diverse populations, as with the COVID-19 vaccine, previously undetected signals (possible associations between vaccines and adverse events that warrant further investigation) may become apparent [7].

The detection of such signals has traditionally been done via passive surveillance (spontaneous reporting systems) because they are somewhat inexpensive [8]. Some downsides, however, are they require health care practitioners to submit reports and are limited by underreporting or selective reporting [8]. A recently emerging alternative for generating vaccine safety evidence is through the use of electronic health records (EHRs), claims, and other available data. This approach has been divided into signal generation and signal evaluation [9], which are conducted using epidemiological designs such as historical comparator and self-controlled case series (SCCS). Whereas signal generation uses a highly sensitive but not specific method that can quickly generate hypotheses about signals, signal evaluation involves a complex and computationally expensive method that is highly specific but not sensitive. Monitoring programs that use a two-stage approach include the Vaccine Safety Datalink and the FDA Center for Medicare & Medicaid Services (CMS). It is becoming difficult to distinguish between these two methods (one simple and one complex) as new softwares and analytic techniques are rapidly developed to investigate vaccine safety signals.

In this study, we evaluated whether the serial strategy (first applying a sensitive method and then a specific method) is useful for signal detection. Epidemiologists commonly use this method for exposure and outcome classification [10][11]. A “gold standard” serial testing method would have 100% sensitivity and specificity [12]. Prior studies have analyzed the statistical accuracy of combining diagnostic tests [13][14]; however, they are focused on understanding tests for patient-level screening, rather than population-level signals for surveillance, which are of interest here. Understanding signal detection for surveillance is relatively uncommon and new.

It remains unknown whether and how using a serial strategy in vaccine surveillance based on available data improves the overall sensitivity and specificity of signal detection and validation, and therefore generates fewer false positive and false negative population-level signals. Specifically, we evaluated performance metrics when the historical comparator design (shown elsewhere to be relatively sensitive) [15] was followed by SCCS (shown elsewhere to be relatively specific) [16], using negative and imputed positive control outcomes in patients receiving historic vaccines across four national administrative claims and EHR databases [17].

## 2 Materials and Methods

### 2.1 Databases

To evaluate the comparative performance of epidemiological designs, we obtained de-identified clinical records from four large administrative claims and EHR databases that used the Observational Medical Outcomes Partnership (OMOP) common data model (CDM) in the United States: IBM MarketScan Commercial Claims and Encounters (CCAE), IBM MarketScan Medicare Supplemental Database (MDCR), IBM MarketScan Multi-State Medicaid Database (MDCD), and Optum® de-identified Electronic Health Record dataset (Optum EHR).

Of the four sources, three (IBM CCAE, IBM MDCR, and IBM MDCD) included adjudicated health insurance claims for commercially insured individuals younger than 65 years old, commercially insured individuals 65 years or older, and racially diverse Medicaid enrollees, respectively. Optum EHR contained electronic health records, covering the general US population. Please refer to Table 1 for additional details.

**Table 1:** Three administrative claims and one EHR data sources with details on population, patients, and history

| Data Source | Population | Patients | History | Description |
| --- | --- | --- | --- | --- |
| IBM MarketScan Commercial Claims and Encounters Database (CCAE) | Commercially insured, <65 years | 142M | 2000- | Enrollees in US employer-sponsored insurance health plans. Adjudicated health insurance claims (e.g. inpatient, outpatient, and outpatient pharmacy), lab tests, and enrollment data from large employers and health plans who provide private healthcare coverage. Includes various fee-for-service, preferred provider organizations, and capitated health plans. |
| IBM Health MarketScan® Multi-State Medicaid Database (MDCD) | Medicaid enrollees, racially diverse | 26M | 2006- | Adjudicated US health insurance claims for Medicaid enrollees. Contains hospital discharge diagnoses, outpatient diagnoses and procedures, and outpatient pharmacy claims. Lacks lab data. |
| IBM MarketScan Medicare Supplemental and Coordination of Benefits Database (MDCR) | Commercially insured, 65+ years | 10M | 2000- | Health services of retirees in US with primary or Medicare supplemental coverage through privately insured fee-for-service, point-of-service, or capitated health plans. Adjudicated health insurance claims (e.g. inpatient, outpatient, and outpatient pharmacy). Lab tests for some. |
| Optum de-identified Electronic Health Record Dataset (Optum EHR) | US, general | 93M | 2006- | Derived from dozens of healthcare provider organizations in US (700+ hospitals and 7,000+ clinics treating 103+ million patients). Clinical information, prescriptions, lab results, vital signs, body measurements, diagnoses, procedures, and information derived from clinical Notes using Natural Language Processing. |

### 2.2 Exposures of interest

We selected six historic vaccine exposures of interest: hemagglutinin type 1 / neuraminidase type 1 (2009 pandemic influenza) (H1N1pdm), seasonal influenza (Fluvirin), seasonal influenza (Fluzone), seasonal influenza (All), zoster (Shingrix; first or second dose), and human papillomavirus (HPV) (Gardasil 9; first or second dose). Each exposure was evaluated over a specific, historical, one-year start and end date (see Table 2).

**Table 2:** Six historic (groups of) vaccines evaluated over specific one-year start and end dates

| Exposure Name | Shot | Start Date | End Date | History Start Date | History End Date |
| --- | --- | --- | --- | --- | --- |
| H1N1 vaccination | First | Sep 1, 2009 | May 31, 2010 | Sep 1, 2008 | May 31, 2009 |
| Seasonal flu vaccination (Fluvirin) | First | Sep 1, 2017 | May 31, 2018 | Sep 1, 2016 | May 31, 2017 |
| Seasonal flu vaccination (Fluzone) | First | Sep 1, 2017 | May 31, 2018 | Sep 1, 2016 | May 31, 2017 |
| Seasonal flu vaccination (All) | First | Sep 1, 2017 | May 31, 2018 | Sep 1, 2016 | May 31, 2017 |
| Zoster vaccination (Shingrix) | Both | Jan 1, 2018 | Dec 31, 2018 | Jan 1, 2017 | Dec 31, 2017 |
| HPV vaccination (Gardasil 9) | Both | Jan 1, 2018 | Dec 31, 2018 | Jan 1, 2017 | Dec 31, 2017 |

*Refer to Appendix A of the study protocol for the formal cohort definitions of each exposure.*

### 2.3 Negative control outcomes

We generated a single list of 93 negative control outcomes (see Supplementary Table 1), which were outcomes not believed to be caused by any vaccines of interest. To identify negative control outcomes that match the severity and prevalence of suspected vaccine adverse effects, we generated a candidate list of negative controls based on similarity of prevalence and percent of diagnoses that were recorded in an inpatient setting (as a proxy for severity). Clinicians reviewed the list to confirm their negative status. Effect size estimates for negative control outcomes should be close to the null; a detected signal away from the null would indicate Type I error.

### 2.4 Positive control outcomes

We modified the 93 negative controls to generate 279 imputed positive control outcomes (outcomes known to be caused by the vaccines) by multiplying the estimated effect size of each negative control by 1.5, 2, and 4. A uniform effect size was used to compute estimates before either of the designs (historical comparator or SCCS) were applied. We chose imputed positive controls instead of real positive controls because well-established vaccine adverse events are rare or carefully monitored such that real-world data do not clearly convey the risk and magnitude of their associations. We used the imputed positive controls to evaluate Type II errors.

### 2.5 Choice of epidemiological designs

To assess the hypothesis that serial testing is favorable, we chose one epidemiological design suspected to be highly sensitive but not highly specific (historical comparator) [16]. Historical comparator compares the observed incidence of adverse events following immunization (AEFI) with the expected incidence of AEFI, estimated from an unexposed patient population, often before or “historical” to vaccine introduction. We adjusted for age and sex, as recommended by a previous study [18], and used a time-at-risk of 1-28 days after the historic visit (defined as a random outpatient visit) for both first and second doses. Any outcomes during that time were attributed to the vaccine [18]. Early signal detection during the historical comparator method can be done if some adverse events are present; however, this can also introduce confounding or false/missed signaling if background rates are inexact or change over time [19]. Then, we selected one design that was suspected to be specific but not sensitive (SCCS) [16]. SCCS compares the time shortly following vaccination to all other time in the same patient’s record, therefore focusing our study on immediate or short-term adverse events [20] [21]. We used a SCCS design adjusted for age and season and excluded a 30-day pre-vaccination window from the analyses to account for healthy vaccinee bias [22].

### 2.6 Performance metrics

We computed effect size estimates with 95% confidence intervals (CIs) and one-sided P values across all databases and all exposures for the historical comparator method alone, SCCS method alone, and the methods combined (historical comparator followed by SCCS). We distinguished signals from non-signals by using P < 0.05 when both designs were applied separately, as well as when they were applied serially (with SCCS applied to signals generated by the historical comparator).

A primary concern in observational studies is the presence of systematic error that may exist because exposed and unexposed populations are not experimentally randomized, but simply observed [23]. Unlike random error, systematic error does not approach zero by merely increasing sample size [24]. It is therefore relatively more problematic when using large databases, such as in our study that captures records for millions of patients. Empirical calibration is a statistical procedure used to adjust for such systematic error [25]. It derives a null distribution from a sample of negative control outcomes and then applies the distribution to unknown effect size estimates, calibrating P values so that 5% of negative controls have P < 0.05 [24]. We can similarly calibrate 95% CIs by modifying negative controls to synthesize positive controls. After calibration, the coverage for a 95% CI is closer to the expected 95% [24]. Without calibration, certain biases may be left unaccounted for [25]. One important assumption of empirical calibration is that the systematic error of the exposure-outcome pair of interest draws from the same distribution as the systematic error for the negative control. Even though it does not require any negative control to have the exact same confounding structure as the exposure-outcome of interest, a weaker assumption of exchangeability is still required. Here we use P-value calibration and make no assumption about systematic error when the true effect size differs from the null.

We analyzed Type I and II errors before and after empirical calibration. A tradeoff exists between the two errors, leading to an increase in Type II error depending on how much systematic error is adjusted for.

Please refer <u>here</u> to access the protocol and analytic code.

## 3 Results

### 3.1 Type I and II errors before empirical calibration for all databases

Figure 1 reports Type I and II error rates in classifying positive and negative control outcomes across vaccine exposure groups. We further compare historical comparator and SCCS with the serial combination of those designs on the y-axis. Bars to the left of 0 indicate Type I error; bars to the right of 0 indicate Type II error. Type I error rate has a nominal cutoff of 0.05 as shown by the dotted line to the left of 0. Error bars closer to 0 indicate higher sensitivity and specificity.

**Figure 1.**
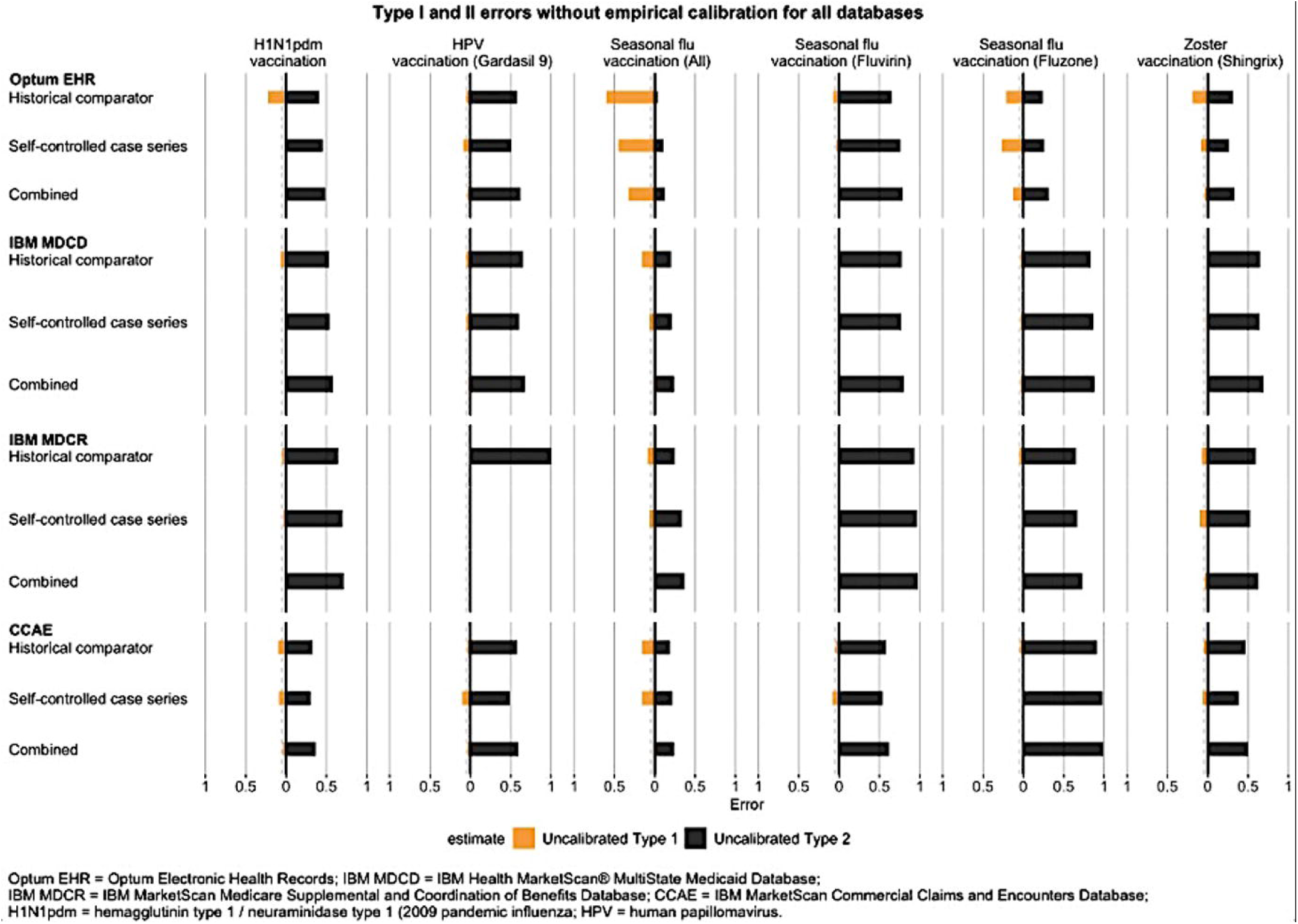
We depict uncalibrated Type I and II errors across all databases using all exposures and negative and positive controls during a time-at-risk of 1-28 days following vaccination. Generally, the historical comparator had low Type II error. SCCS had low Type I error. Clinical intuition would predict the combined method to be highly sensitive and specific, but results showed high specificity and low sensitivity.

Almost all analyses had Type II error rates exceeding 50% (Figure 1). Meanwhile, Type I error bars tended to range between 0% and 50%, except for seasonal influenza (All) in the Optum EHR database where the historical comparator method generated many false positive signals.

The difference between Type I and II error rates before empirical calibration was particularly visible for the Optum EHR database. In most scenarios, the historical comparator method had higher sensitivity than SCCS or combined. When the historical comparator design was used to identify adverse events following vaccination, it generated fewer false negative signals (was less likely to miss a signal when it existed). Meanwhile, SCCS was more specific than the historical comparator method for H1N1pdm, seasonal influenza (All), seasonal influenza (Fluvirin), and zoster (Shingrix). The serial approach generated more false negative signals, while often reducing the number of false positives. This was consistent across all four databases: serial combination did not improve overall performance.

### 3.2 Type I and II errors after empirical calibration for all databases

We also considered how empirical calibration changed Type I and II errors (Figure 2). After calibration, Type I errors returned to nominal, and Type II errors increased in most cases. For the Optum EHR database, calibration visibly reduced Type I error rates of the historical comparator method for seasonal influenza (All). In spite of this, the combined method did not improve overall performance in the way that our hypothesis predicted. Among all three design choices and databases, sensitivity was lowest when the methods were serially combined (it increased the number of times that the surveillance system missed a signal when it actually existed). Yet, this may be expected since only signals flagged in the historical comparator method are then evaluated in SCCS, so there is an opportunity to decrease false positive signals.

**Figure 2.**
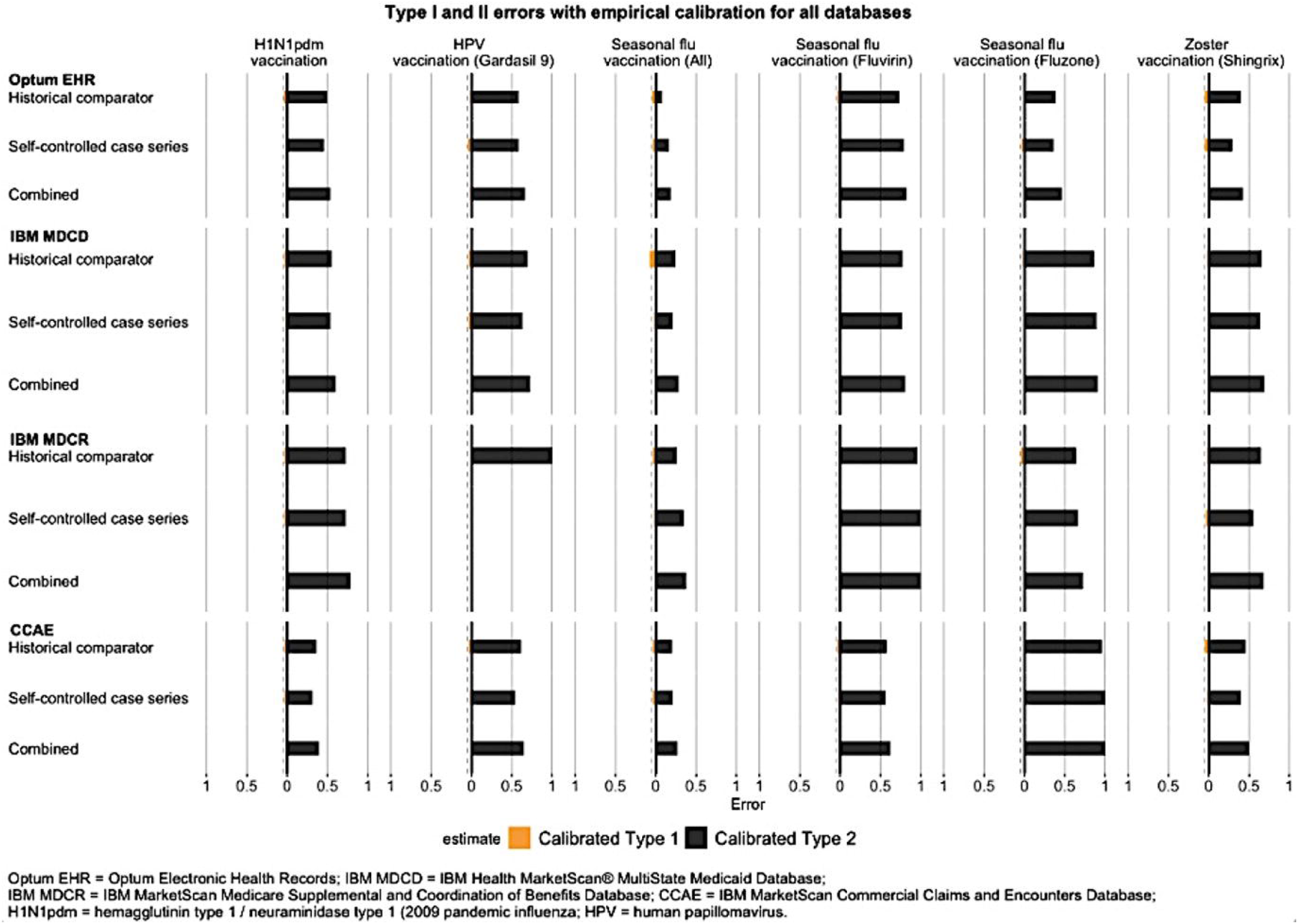
We depict Type I and II errors after empirical calibration across all databases using all exposures and negative and positive controls during a time-at-risk of 1-28 days following vaccination. Type I errors returned to nominal. Type II errors mostly increased. Even with empirical calibration, the serial approach did not increase overall performance of a two-stage surveillance program.

## 4 Discussion

### 4.1 Key results

In this large-scale observational study of vaccine surveillance using four real-world databases in the US, we found advantages and disadvantages to serially combining sensitive and specific methods. We reject our hypothesis that combining designs increases sensitivity and specificity. Serial combination was helpful because it showed reduced Type I error, but also flawed because Type II error increased to be higher than when either of the methods were used alone. This tradeoff in Type I and II error is expected. Only signals flagged in the historical comparator were evaluated in the second method SCCS, creating an opportunity to remove false positive signals but not false negative signals. Also, SCCS typically already had quite favorable Type I error rates when applied individually, so combining methods did little to further decrease Type I error. Because our results showed false negative signals were excessively higher than false positive signals, the notion of dividing surveillance into two distinct stages of signal generation and evaluation, might not be ideal. We did not calculate minimum detectable relative risk (MDRR).

Although preferences for sensitivity and specificity trade-off may vary, an increase in specificity demonstrably did not offset a decrease in sensitivity. If specificity is high, but sensitivity is low, potential adverse events that might be caused by a vaccine can go overlooked, allowing for further circulation of the vaccine in the market. If sensitivity is high and specificity is low, adverse events might be perceived as associated with a vaccine when in truth they are not, leading to the withdrawal of a vaccine from the market. In either case, an error has been made. Sensitivity alone does not predict enough about negative signals, and specificity alone does not predict enough about positive signals (“worst of both worlds”).

Other studies have also examined performance characteristics in serial monitoring. Nelson et al. (2012) studied the use of serial approaches with observational EHR data in identifying signals between the DTap-IPV-Hib vaccine and adverse events [26]. One analytic challenge was confounding introduced from differences in vaccine uptake by age group [26]. Since observational studies are inherently uncontrolled, we corrected for confounding via empirical calibration.

Prior research has suggested that the historical comparator method is highly sensitive. For example, Li et al. recently found that historical comparisons had Type II errors ranging between 0% to 10% and Type I errors above 30% [18]. Age and sex adjustment reduced Type I error, had little impact on Type II error, and improved precision in some cases [16]. Historical rate comparison was, overall, good at identifying true safety signals [16]. Based on Li et al.’s results, we evaluated the age and sex adjusted variant for historical comparator, using a time-at-risk of 1-28 days after the historic visit, for both first and second doses. However, contrary to this belief, we observed some instances where SCCS had lower Type II error and was therefore more sensitive than the historical comparator method. For example, in Figure 1, this was present for outcomes involving HPV (Gardasil 9) and zoster (Shingrix) in the Optum EHR and CCAE databases. There was also almost no distinguishable difference between the Type II errors for the historical comparator and SCCS methods in several of the plots, including for H1N1pdm and seasonal influenza (Fluvirin) in IBM MDCD, as well as seasonal influenza (Fluzone) in IBM MDCR. Likewise, there were some incidences where SCCS was less specific than historical comparator, such as in data for seasonal influenza (Fluzone) in Optum EHR, zoster (Shingrix) in IBM MDCR, and HPV (Gardasil 9) in CCAE. Comparisons of performance metrics demonstrate that the historical comparator is not always most sensitive. This uncertainty can make it difficult to predict the utility of a serial approach. Finally, Maro et al. (2014) conducted a similar study on sequential database surveillance using first an unmatched historical comparison method and second a self-controlled risk interval design (SCRI) [27]. Their time-at-risk was 1-42 days. However, this study implemented both methods using a sequential framework instead of serial approach and provided limited information on the sensitivity and specificity of combining methods.

On the individual patient-level scale, serial approach is common worldwide in the realm of diagnostic testing. Consider, for instance, detecting human immunodeficiency virus (HIV) prevalence in a population. A study amongst Nigerian women found that serial rapid tests (where the first HIV test has a sensitivity close to 100%, and the second test is implemented only if the initial results were positive) were more sensitive and specific than conventional HIV-testing techniques [28]. Similar studies that used HIV rapid serological testing using data from Cote d’Ivoire [29], Uganda [30], and India [31] found that serial testing provides reliable results and has benefits in resource-limited areas. Serial testing has been widely adopted in clinical screenings for tuberculosis [32], prostate cancer [33], acute coronary syndrome [34], ovarian cancer [35], and recently COVID-19 [36]. This principle of investigating diseases using a cheap method before an expensive one is central to clinical medicine.

The importance of our study for the future of safety surveillance is related to recommended measures that should be addressed before using observational electronic healthcare databases for safety surveillance [37], namely that of pre-specifying and evaluating statistical designs. An accurate epidemiological design should overall generate the fewest number of false positive and false negative signals. Combining designs is unlikely to achieve this goal. Individuals who prioritize specificity over sensitivity might claim serial combination did work. Further studies should examine when this tradeoff is worthwhile. Since we observed variability in results, there should be similar analyses with other databases and exposures, such as COVID-19 vaccines or populations for which a certain occurrence has been defined (e.g. pregnant women or those with common comorbidities).

### 4.2 Strengths and limitations

Three strengths of this work are the use of real-world data, mapping of data to the OMOP CDM, and open-source availability of our code. First, real-world data represents a more heterogeneous population than randomized controlled trials that are tightly controlled. This improves the external validity of our data to the real-world patient population. Second, the CDM unifies data from multiple partners into a standardized format and enables us to pool analysis results from sources with large numbers of study subjects, without having to expose patient-level information. Third, the open-source availability of our code and protocol enables transparency and collaboration.

Our study is limited by the vaccine exposure of interest. While we seek to understand the use of epidemiological designs not only in general but also for COVID-19 vaccine surveillance, none of our exposures were COVID-19 vaccines. We addressed this limitation by choosing historical viral vaccines, which may be most similar. Another limitation is the use of imputed positive controls that assumes that systematic error does not change as a function of true effect size.

While Vaccine Safety Datalink and FDA CMS use serial combination with the best of intent, it may be the worst of both worlds (sensitivity and specificity). As COVID-19 vaccine safety surveillance becomes increasingly important, efforts to implement monitoring systems should carefully consider the utility and sequence of epidemiological designs.

## Data Availability

The protocol and analytical code underlying this manuscript are available in the Eumaeus Repository at https://github.com/ohdsi-studies/Eumaeus.

## 6 Ethics Statement

This study did not require Institutional Review Board approval from data partners because it used de-identified patient data and did not constitute research with human subjects.

## 7 Author Contributions

**FA, MJS, PBR, GH, DPA** and **MAS** conceived the research. **FA, MJS** and **MAS** drafted the manuscript in consultation with **EPM, TMA, LYHL, TDS, SF, FN, PBR, GH, DPA** who provided critical feedback on the research and writing. All authors read and approved the final version.

## 10 Acknowledgments

We are indebted to the entire EUMAEUS task force within the OHDSI community.

