## Supplementary Table for "Best of intent, worst of both worlds: why sequentially combining epidemiological designs does not improve signal detection in vaccine safety surveillance"

| Outcome Id | Outcome Name |
|---|---|
| 438945 | Accidental poisoning by benzodiazepine-based tranquilizer |
| 434455 | Acquired claw toes |
| 316211 | Acquired spondylolisthesis |
| 201612 | Alcoholic liver damage |
| 438730 | Alkalosis |
| 441258 | Anemia in neoplastic disease |
| 432513 | Animal bite wound |
| 4171556 | Ankle ulcer |
| 4098292 | Antiphospholipid syndrome |
| 77650 | Aseptic necrosis of bone |
| 4239873 | Benign neoplasm of ciliary body |
| 23731 | Benign neoplasm of larynx |
| 199764 | Benign neoplasm of ovary |
| 195500 | Benign neoplasm of uterus |
| 4145627 | Biliary calculus |
| 4108471 | Burn of digit of hand |
| 75121 | Burn of lower leg |
| 4284982 | Calculus of bile duct without obstruction |
| 434327 | Cannabis abuse |
| 78497 | Cellulitis and abscess of toe |
| 4001454 | Cervical spine ankylosis |
| 4068241 | Chronic instability of knee |
| 195596 | Chronic pancreatitis |
| 4206338 | Chronic salpingitis |
| 4058397 | Claustrophobia |
| 74816 | Contusion of toe |
| 73302 | Curvature of spine |
| 4151134 | Cyst of pancreas |
| 77638 | Displacement of intervertebral disc without myelopathy |
| 195864 | Diverticulum of bladder |
| 201346 | Edema of penis |
| 200461 | Endometriosis of uterus |
| 377877 | Esotropia |
| 193530 | Follicular cyst of ovary |
| 4094822 | Foreign body in respiratory tract |
| 443421 | Gallbladder and bile duct calculi |
| 4299408 | Gouty tophus |
| 135215 | Hashimoto thyroiditis |
| 442190 | Hemorrhage of colon |
| 43020475 | High risk heterosexual behavior |
| 194149 | Hirschsprung’s disease |
| 443204 | Human ehrlichiosis |
| 4226238 | Hyperosmolar coma due to diabetes mellitus |
| 4032787 | Hyperosmolarity |
| 197032 | Hyperplasia of prostate |

| | |
|---|---|
| 140362 | Hypoparathyroidism |
| 435371 | Hypothermia |
| 138690 | Infestation by Pediculus |
| 4152376 | Intentional self poisoning |
| 192953 | Intestinal adhesions with obstruction |
| 196347 | Intestinal parasitism |
| 137977 | Jaundice |
| 317510 | Leukemia |
| 765053 | Lump in right breast |
| 378165 | Nystagmus |
| 434085 | Obstruction of duodenum |
| 4147016 | Open wound of buttock |
| 4129404 | Open wound of upper arm |
| 438120 | Opioid dependence |
| 75924 | Osteodystrophy |
| 432594 | Osteomalacia |
| 30365 | Panhypopituitarism |
| 4108371 | Peripheral gangrene |
| 440367 | Plasmacytosis |
| 439233 | Poisoning by antidiabetic agent |
| 442149 | Poisoning by bee sting |
| 4314086 | Poisoning due to sting of ant |
| 4147660 | Postural kyphosis |
| 434319 | Premature ejaculation |
| 199754 | Primary malignant neoplasm of pancreas |
| 4311499 | Primary malignant neoplasm of respiratory tract |
| 436635 | Primary malignant neoplasm of sigmoid colon |
| 196044 | Primary malignant neoplasm of stomach |
| 433716 | Primary malignant neoplasm of testis |
| 133424 | Primary malignant neoplasm of thyroid gland |
| 194997 | Prostatitis |
| 80286 | Prosthetic joint loosening |
| 443274 | Psychostimulant dependence |
| 314962 | Raynaud’s disease |
| 37018294 | Residual osteitis |
| 4288241 | Salmonella enterica subspecies arizonae infection |
| 45757269 | Sclerosing mesenteritis |
| 74722 | Secondary localized osteoarthrosis of pelvic region |
| 200348 | Secondary malignant neoplasm of large intestine |
| 43020446 | Sedative withdrawal |
| 74194 | Sprain of spinal ligament |
| 4194207 | Tailor’s bunion |
| 193521 | Tropical sprue |
| 40482801 | Type II diabetes mellitus uncontrolled |
| 74719 | Ulcer of foot |
| 196625 | Viral hepatitis A without hepatic coma |
| 197494 | Viral hepatitis C |
| 4284533 | Vitamin D-dependent rickets |
